# A Modified Percutaneous Spinal Cord Stimulation Implant Approach to Target the Ventral Spinal Cord

**DOI:** 10.64898/2026.04.06.26350176

**Authors:** Kyle J. Valestrino, Chimdi V. Ihediwa, Geoffrey T. Dorius, Aaron M. Conger, Allison Glinka-Przybysz, Zachary L. McCormick, Alexandra E. Fogarty, Mark A. Mahan, Jose Hernandez-Bello, Peter E. Konrad, Taylor R. Burnham, Ashley N. Dalrymple

## Abstract

**Objectives:** Epidural spinal cord stimulation (SCS) is an emerging therapy for motor rehabilitation following spinal cord injury (SCI) and other motor disorders. Conventionally, SCS leads are placed along the dorsal spinal cord (SCS_D_), where stimulation activates large diameter afferent fibers, which indirectly activate motoneurons through reflex pathways. This leads to broad activation of flexor and extensor muscles and limited fine-tuned control of motor output. Targeting the ventral spinal cord (SCS_V_) may enable more direct activation of motoneuron pools, potentially improving the specificity of muscle activation; however, there is currently no established method to place leads ventrally. To address this, we evaluated the feasibility of four modified percutaneous implantation techniques to target the ventrolateral thoracolumbar spinal cord.

**Materials and methods:** Percutaneous SCS_V_ implantation was performed in three human cadaver torso specimens under fluoroscopic guidance. The following approaches were evaluated: sacral hiatus, transforaminal, interlaminar contralateral, and interlaminar ipsilateral. The leads in the latter 3 approaches were inserted between L1 and L5. Eighteen implants were attempted, with nine leads retained for analysis. Lead and electrode position were assessed using computed tomography (CT) with three-dimensional reconstruction, along with anatomical dissection to verify lead and electrode placement within the epidural space.

**Results:** Successful ventral epidural lead placement was achieved using all four implantation approaches. The sacral hiatus (16/16 electrodes) and transforaminal (8/8 electrodes) approaches resulted in exclusively ventrolateral placement. The interlaminar contralateral approach led to 27/32 electrodes positioned ventrolaterally and 5/32 dorsally. The interlaminar ipsilateral implantation approach led to 14/32 electrodes positioned ventrolaterally and 18/32 positioned ventromedially.

**Conclusions:** These findings demonstrate that ventral epidural SCS lead placement can be achieved using modified percutaneous implant techniques. The four approaches outlined here provide a clinically feasible pathway to SCS_V_ and establishes a foundation for future clinical studies investigating SCS_V_ for motor rehabilitation following SCI.

## Introduction

Spinal cord injury (SCI) is a prevalent and life-altering condition, affecting roughly 20.6 million people worldwide (1). Individuals with SCI often experience permanent loss of partial or full motor function, resulting in significantly reduced quality of life and independence (1). Despite advances in acute care and rehabilitation training, functional recovery after severe paralysis remains limited (2). Currently, the standard of care for those with severe SCI relies largely on compensatory strategies, and meaningful neurological recovery is rare once chronic paralysis is established (3,4). There is, therefore, a critical need for new therapeutic strategies to restore motor function in this population. One promising approach is spinal cord stimulation (SCS), a neuromodulation therapy that has helped improve rehabilitation outcomes for patients with chronic paralysis of either upper or lower extremities (5).

SCS is an established clinical intervention for chronic neuropathic pain that is refractory to other treatments (6). In conventional SCS, electrodes are placed epidurally over the dorsal aspect of the spinal cord and dorsal roots (SCS_D_; Figure 1), where they activate large-diameter afferent fibers to inhibit pain transmission via the gate control theory and other mechanisms (6– 8). Recently, a number of clinical research studies have demonstrated that SCS_D_ can enhance motor rehabilitation after chronic SCI (5,9–11). These studies have shown promising results in functional recovery, demonstrating that SCS applied over the lumbosacral spinal cord can evoke motor responses in lower limb musculature (9,12–14), as well as enable voluntary movements in individuals with complete paralysis (11,15–17).

**Figure 1.**
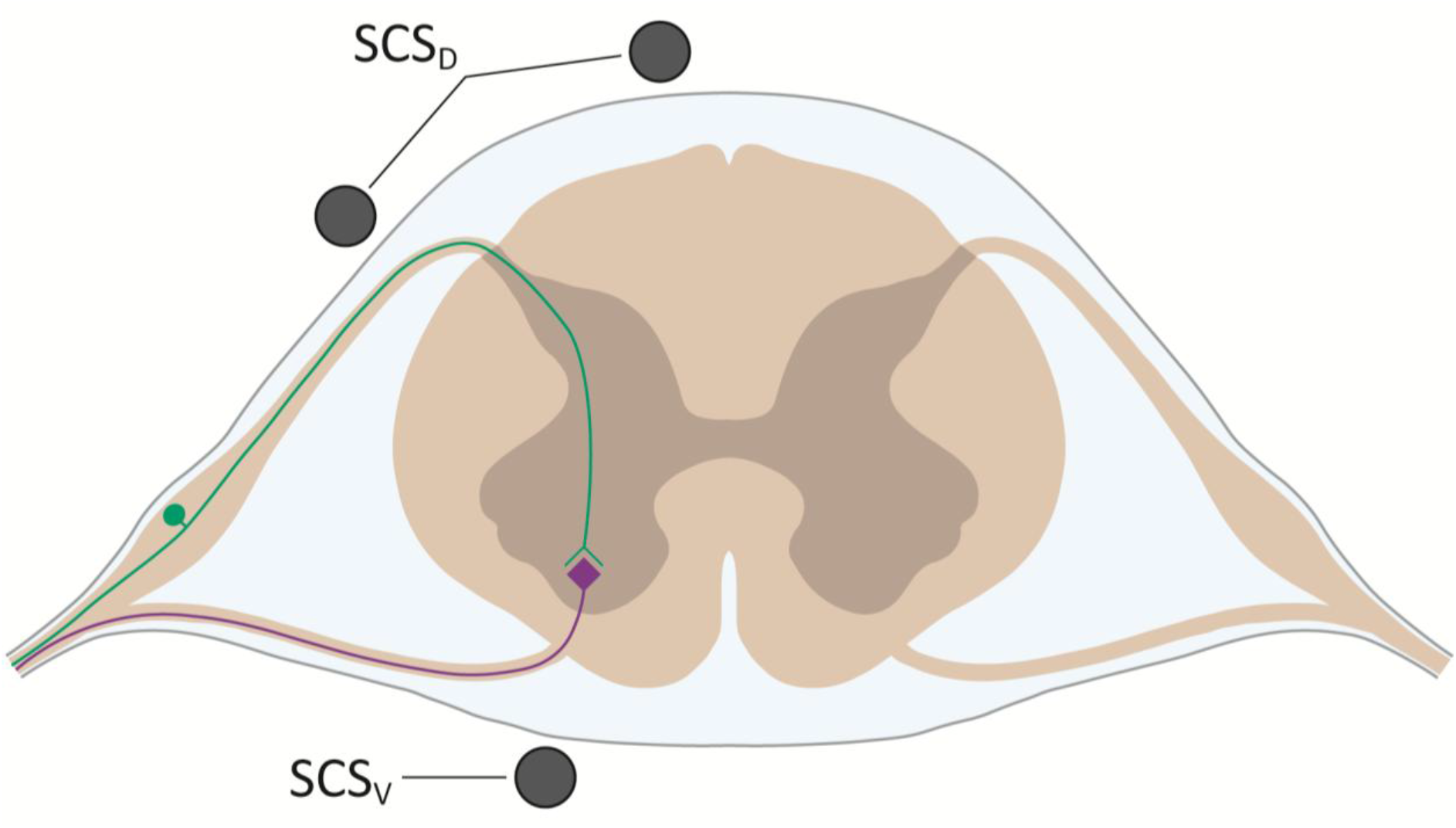
Ventral and dorsal epidural SCS. Cross-sectional schematic showing dorsal (SCS_D_) and ventral (SCS_V_) epidural lead placement relative to the spinal cord. SCS_D_ is located closer to afferent fibers (green) in the dorsal roots as well as the dorsal column, while SCS_V_ is placed closer to the motoneurons in the ventral horn (purple).

Despite these promising results, SCS_D_ for motor rehabilitation has limitations to consider. Specifically, the conventional dorsal placement of epidural electrodes primarily activates large-diameter afferent fibers, which in turn, indirectly activate motoneurons through reflex pathways (8). As a result, SCS_D_ tends to produce broad flexor extensor movements rather than isolated, selective movements and muscle activation (9,18).

One approach to improve the specificity of epidural SCS is to position the electrodes closer to the motoneuron pools in the ventral horn – a strategy we refer to as ventral spinal cord stimulation (SCS_V_). With SCS_V_, it may be possible to activate motoneurons more directly, thereby eliciting muscle contractions in a more controlled, muscle-specific manner. Preclinical work supports this hypothesis: SCS_V_ has been shown to selectively activate forelimb muscles in a rat model, enabling fine control of motor outputs (19). Similarly, a computational study comparing muscle selectivity of SCS_D_ and SCS_V_ in a model of a rat spinal cord showed that SCS_V_ was more selective (20). Thoracic SCS_V_ has also been performed in humans in an isolated case study (21), demonstrating that access to and stimulation of the ventral surface of the spinal cord is feasible. In that study, SCS_V_ was applied acutely after SCI, producing increased muscle activity and force generation compared to conditions without stimulation. When paired with physical therapy, the participant also showed signs of motor improvement. However, this study relied on a transpedicular corpectomy and laminectomy procedure to access the ventral aspect of the spinal cord (21). As such, a less invasive method for ventral lead placement could help expedite translational research on SCS_V_ as a potential rehabilitation therapy across larger patient cohorts. In this study, we proposed and evaluated a modified percutaneous SCS lead implantation approach to target the ventral spinal cord. The goals of this study were to determine whether ventral epidural lead positioning can be achieved through posterior percutaneous approaches, to characterize the final anatomical placement of these leads relative to the spinal cord and roots, and to identify any technical obstacles or complications. By validating a viable and reliable method for ventral SCS lead placement, this work aims to lay the groundwork for future clinical studies of SCS_V_ as a targeted therapy for motor rehabilitation in people with SCI.

## Methods

### Specimen Information

Three cadaveric torso specimens were obtained from Science Care (Phoenix, AZ, USA). All three were partially embalmed, eviscerated, frozen, and thawed overnight prior to the implant procedure. The specimens included one female, 20-25 years old (Cadaver 1) and two males between 65 and 70 years old (Cadavers 2 and 3). No Institutional Review Board protocol was needed because the study was performed in cadaver specimens.

### Implant Procedure

All implant procedures took place at the Surgical Knowledge Integration (SKI) Lab at the University of Utah. Each was performed with the cadaver positioned prone on a radiolucent table to allow unobstructed fluoroscopic imaging with a GE 9800+ C-Arm fluoroscope (GE Healthcare, Chicago, IL, USA). Three SCS leads were implanted into each cadaver specimen, for a total of nine leads across all procedures. Two Infinion 16 percutaneous leads (Boston Scientific, Marlborough, MA, USA) and seven Nevro Senza percutaneous leads (Nevro, Redwood City, CA, USA) were implanted. Insertions were performed by a team of physicians: four physiatrists (AC, AF, AG, and ZM) and one neurosurgeon (MM).

Under fluoroscopic guidance, a 14-gauge Tuohy epidural needle was used to access the epidural space. Four different insertion techniques were trialed to achieve thoracolumbar ventral placement of the leads (Figure 2):

**Figure 2.**
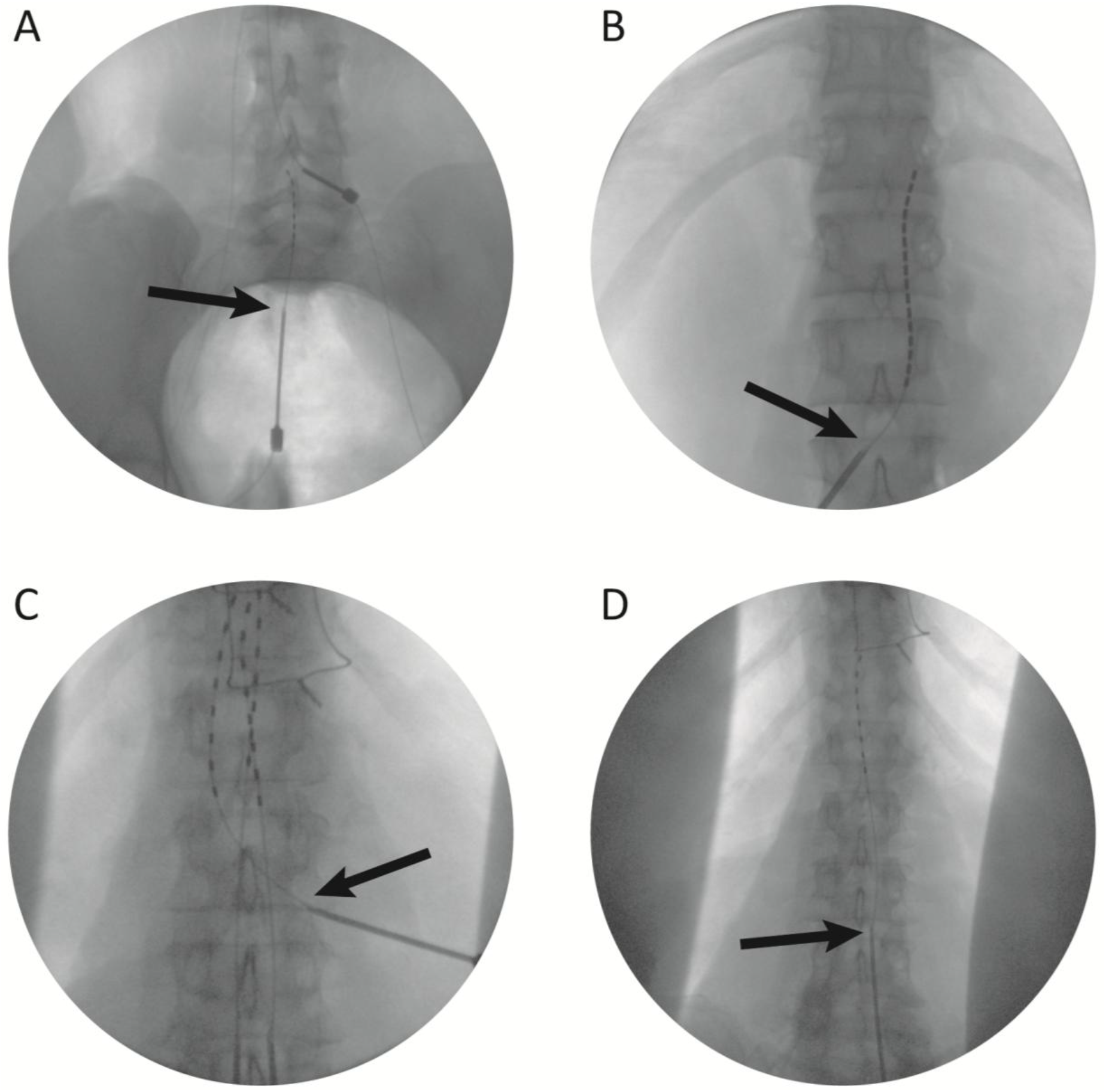
Fluoroscopy images demonstrating the insertion point and trajectory for our four different implant approaches. (A) Sacral hiatus. (B) Transforaminal. (C) Interlaminar contralateral. (D) Interlaminar ipsilateral. Arrows indicate the point of entry into the spinal column.

1. **Sacral hiatus approach:** The needle was introduced through the sacral hiatus (at approximately the S5 level), and the lead was advanced rostrally.
2. **Transforaminal approach:** The needle entered the ventral epidural space directly via the intervertebral foramen and the lead was advanced rostrally.
3. **Interlaminar contralateral approach:** The needle was inserted through the interlaminar space just lateral to midline at an oblique angle relative to the spine, pointing contralaterally. After entering the dorsal epidural space, the lead was directed toward the contralateral side of the spinal canal, around the dural sac, then to the ventral epidural space. Once in the ventral space, the lead was advanced rostrally.
4. **Interlaminar ipsilateral approach:** The needle was inserted through the interlaminar space just lateral to midline. The lead was immediately directed ventrally, around the dural sac, on the ipsilateral side and advanced rostrally.

The levels of insertion and final positioning are shown in Table 1. Across cadavers, multiple insertion attempts were performed to evaluate the feasibility of each approach and for each clinical team member to refine the implant technique. In Cadaver 1, five implants were attempted using the interlaminar contralateral and sacral hiatus approaches. In Cadaver 2, six implants were attempted using interlaminar ipsilateral, transforaminal, and sacral hiatus approaches. In Cadaver 3, seven implants were attempted using both the interlaminar contralateral and interlaminar ipsilateral approaches. Final positioning was confirmed with both anteroposterior and lateral fluoroscopic views. Although multiple implantation attempts were performed, three leads per cadaver were ultimately sutured in place and retained for subsequent computed tomography (CT) imaging and anatomical analysis to determine their final position.

**Table 1.**
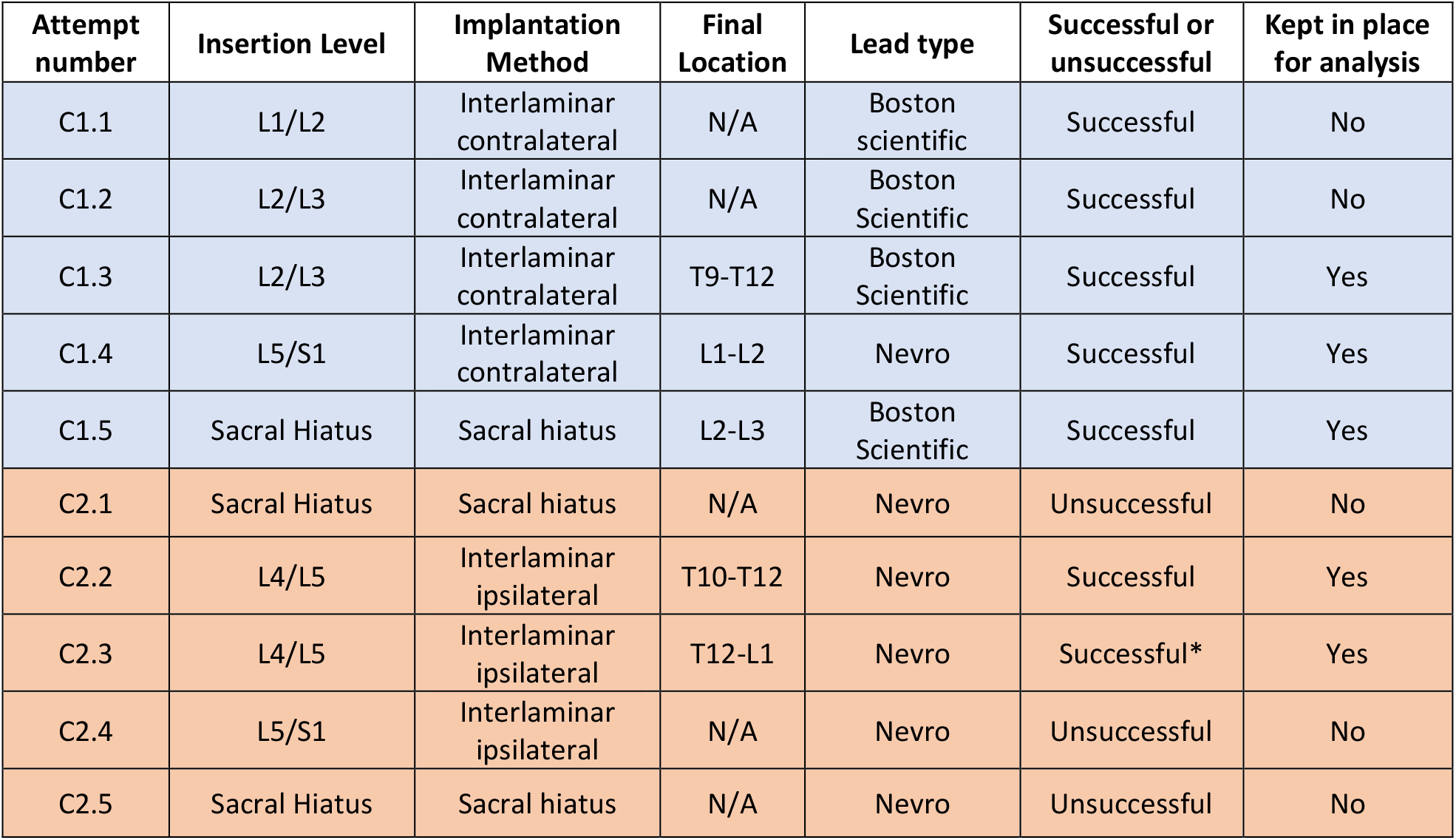
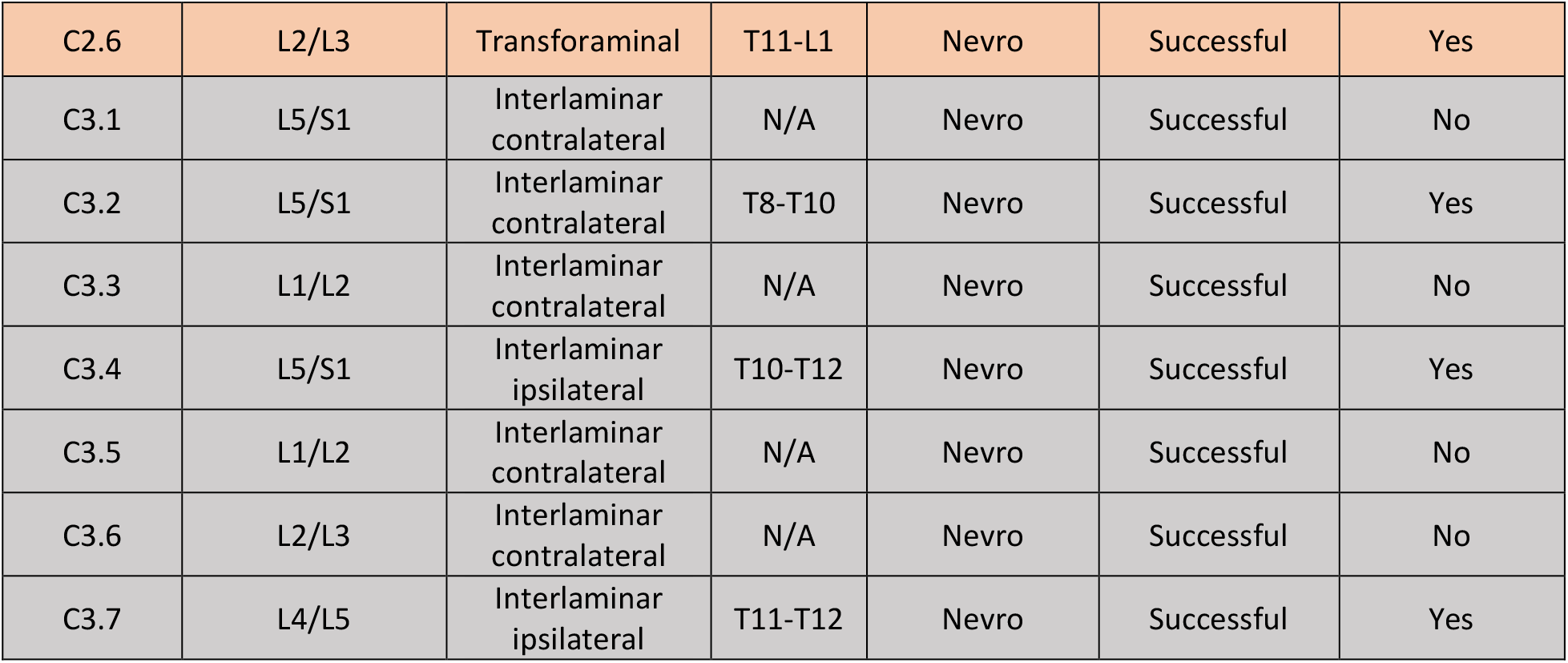
Summary of SCS_V_ electrode locations for all lead insertions attempted. Attempt number includes the cadaver number as C# and are organized by color: Cadaver 1 (blue), cadaver 2 (orange), and cadaver 3 (grey). Implants were defined as successful or unsuccessful based on whether or not we were able to target the ventral epidural space and advance the lead rostrally without complications. Attempt C2.3 appeared successful during the implant, though further analysis revealed that a dural puncture occurred and the lead was inserted subdurally on the ventral side.

| Attempt number | Insertion Level | Implantation Method | Final Location | Lead type | Successful or unsuccessful | Kept in place for analysis |
| --- | --- | --- | --- | --- | --- | --- |
| C1.1 | L1/L2 | Interlaminar contralateral | N/A | Boston scientific | Successful | No |
| C1.2 | L2/L3 | Interlaminar contralateral | N/A | Boston Scientific | Successful | No |
| C1.3 | L2/L3 | Interlaminar contralateral | T9-T12 | Boston Scientific | Successful | Yes |
| C1.4 | L5/S1 | Interlaminar contralateral | L1-L2 | Nevro | Successful | Yes |
| C1.5 | Sacral Hiatus | Sacral hiatus | L2-L3 | Boston Scientific | Successful | Yes |
| C2.1 | Sacral Hiatus | Sacral hiatus | N/A | Nevro | Unsuccessful | No |
| C2.2 | L4/L5 | Interlaminar ipsilateral | T10-T12 | Nevro | Successful | Yes |
| C2.3 | L4/L5 | Interlaminar ipsilateral | T12-L1 | Nevro | Successful* | Yes |
| C2.4 | L5/S1 | Interlaminar ipsilateral | N/A | Nevro | Unsuccessful | No |
| C2.5 | Sacral Hiatus | Sacral hiatus | N/A | Nevro | Unsuccessful | No |
| C2.6 | L2/L3 | Transforaminal | T11-L1 | Nevro | Successful | Yes |
| C3.1 | L5/S1 | Interlaminar contralateral | N/A | Nevro | Successful | No |
| C3.2 | L5/S1 | Interlaminar contralateral | T8-T10 | Nevro | Successful | Yes |
| C3.3 | L1/L2 | Interlaminar contralateral | N/A | Nevro | Successful | No |
| C3.4 | L5/S1 | Interlaminar ipsilateral | T10-T12 | Nevro | Successful | Yes |
| C3.5 | L1/L2 | Interlaminar contralateral | N/A | Nevro | Successful | No |
| C3.6 | L2/L3 | Interlaminar contralateral | N/A | Nevro | Successful | No |
| C3.7 | L4/L5 | Interlaminar ipsilateral | T11-T12 | Nevro | Successful | Yes |

### Computed Tomography

CT imaging was performed using a Siemens SOMATOM Definition Flash scanner (Siemens Healthineers, Erlangen, Germany) spine protocol with a slice thickness of 0.6 mm. DICOM data from each scan were imported into Horos (Horos Project, open-source medical image viewer for macOS, version 3.3.6) for post-processing. A three-dimensional reconstruction was generated for each specimen, and threshold-based voxel intensity filtering was applied to isolate both the spinal column and the implanted SCS_V_ leads. This process enabled clear visualization of lead placement in relation to spinal anatomy for each specimen.

### Anatomical Dissection

Following CT imaging, an anatomical dissection was performed to verify lead placement and assess for gross tissue damage. The spinal levels selected for dissection were based on the final position of the leads, as determined by intraoperative fluoroscopy.

An incision was made along the spine midline, and skin and paraspinal musculature were reflected laterally to expose the spinous processes and laminae. A periosteal elevator was used to remove residual soft tissue from the exposed bone surfaces. A laminectomy was then performed using a bone saw to gain direct access to the spinal canal.

With the dura mater exposed, the dural sac was manipulated to enable visualization of the implanted leads. The position of the leads within the spinal canal was evaluated relative to key neuroanatomical landmarks, such as the nerve roots. The full path of each lead was assessed to confirm accurate placement and to find any signs of damage to surrounding structures.

### Outcome Classification

For each cadaver, three leads were sutured in place for further analysis through CT imaging and anatomical dissection. Using images from the CT scans in the transverse plane, each electrode from all implanted leads were classified as dorsal, ventrolateral, or ventromedial (Figure 3). We analyzed lead location based on the cadaver number as well as the insertion method to determine (1) whether we were able to iteratively improve successful ventral targeting performance across implant sessions and (2) which implantation techniques proved most successful. Unsuccessful and successful implantation attempts that were not sutured in place are not included in the analyses reported below but are outlined in Table 1.

**Figure 3.**
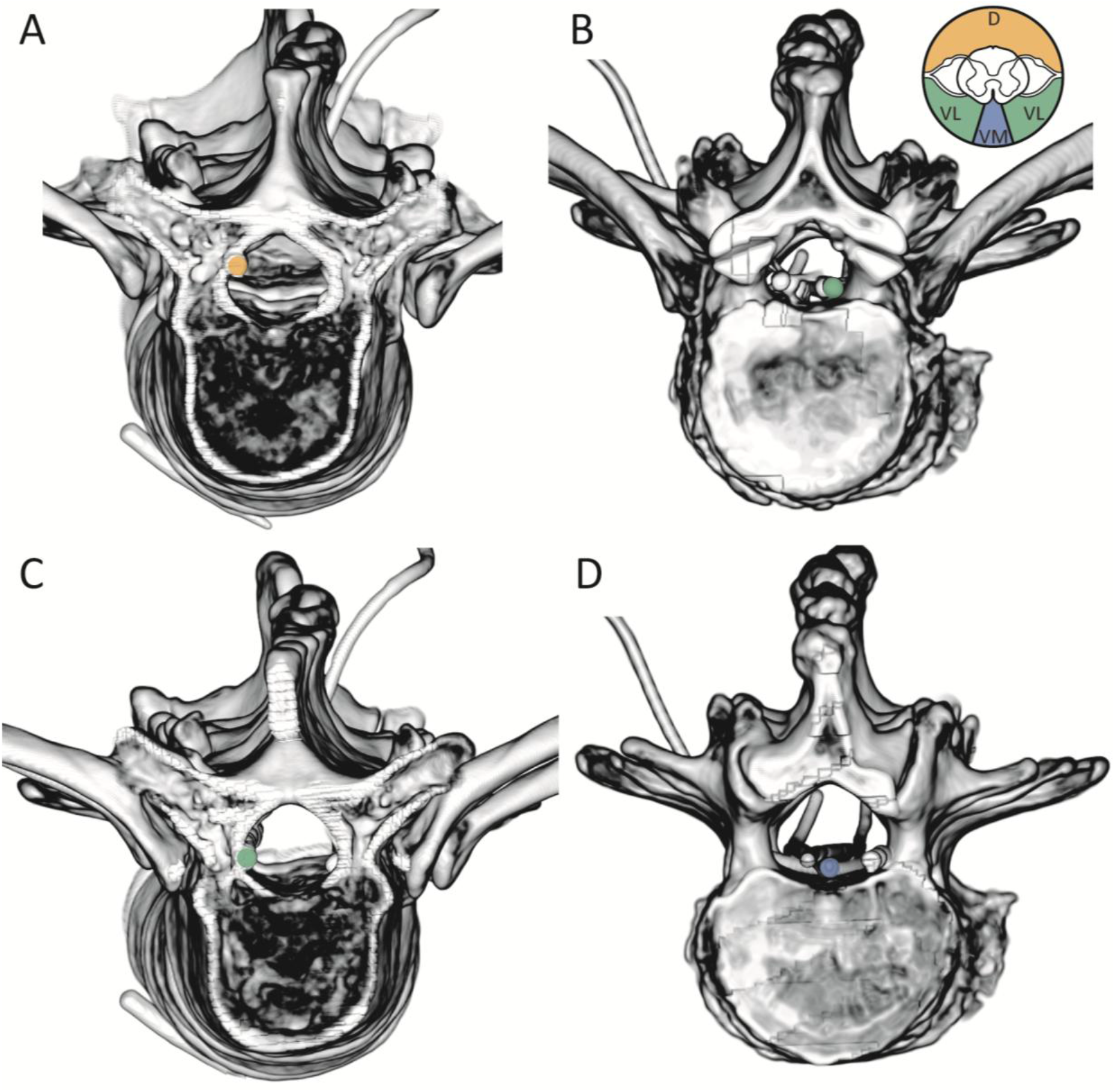
Three-dimensional reconstructions of CT scans were used to classify SCS lead and electrode positions as (A) dorsal (D; yellow), (B), (C) ventrolateral (VL; green), or (D) ventromedial (VM; blue).

## Results

Examples of the lead placement following each of the four implantation approaches in all 3 cadaver specimens are shown in Supplementary Figure 1. A summary of the implant approaches and final position of the electrodes for each cadaver implant section is provided in Table 1 and Figure 4. Representative examples of the electrode–root relationships as viewed during dissection are shown in Figure 5.

**Figure 4.**
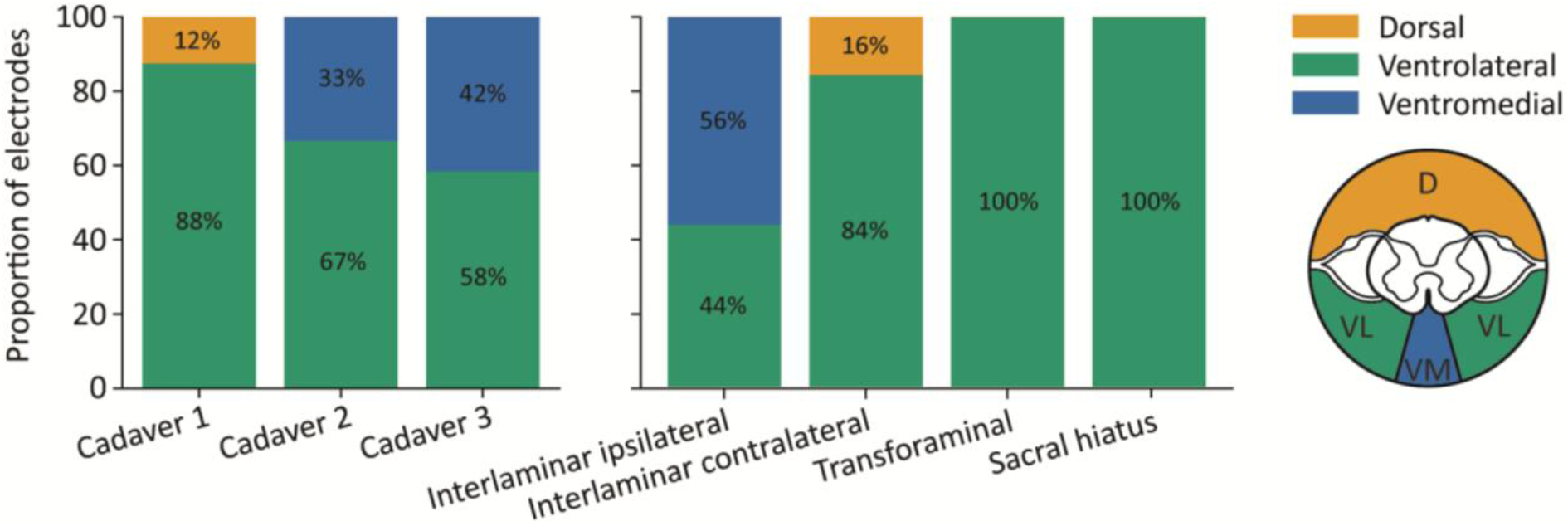
Electrode classification by cadaver (left) and implant approach (right). Stacked bar plots show the proportion of electrodes classified as dorsal, ventromedial, or ventrolateral.

**Figure 5.**
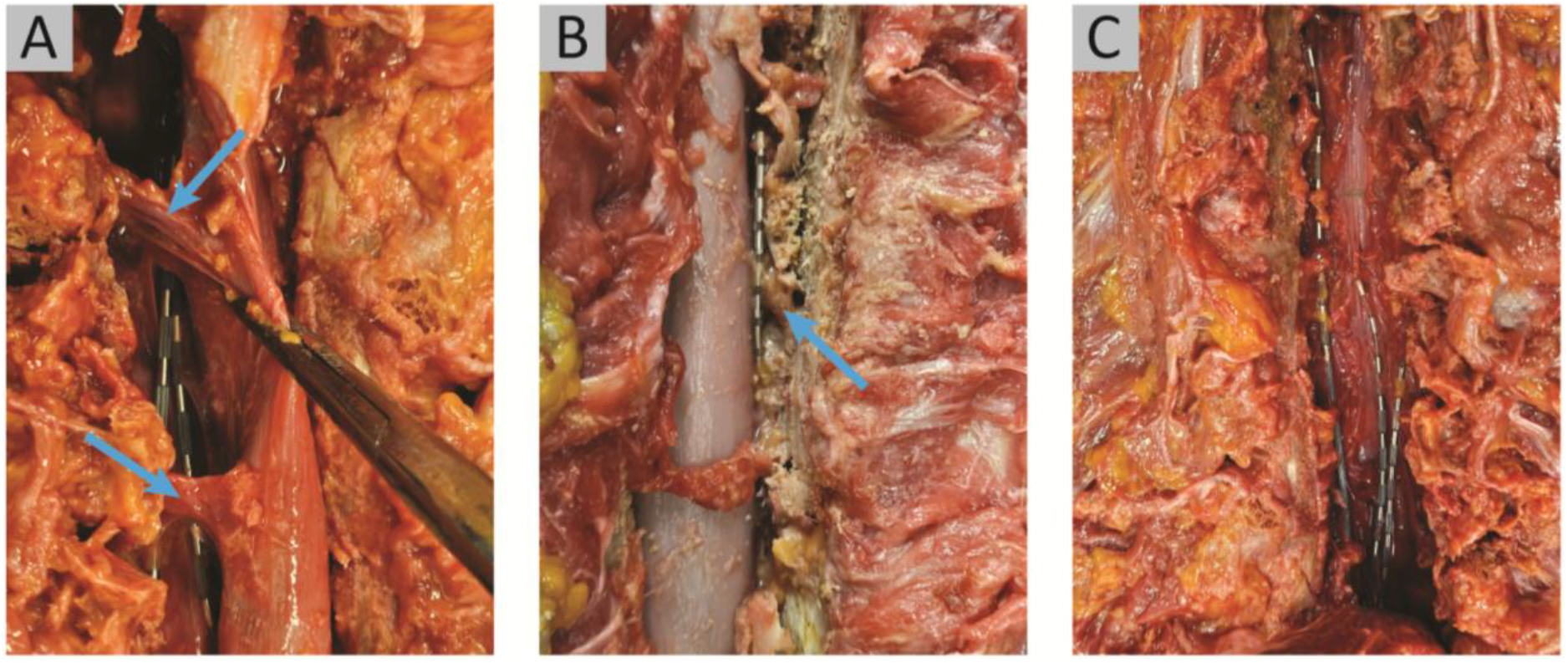
Example images of lead placement as viewed during anatomical dissection. (A) SCS leads positioned ventral to the spinal roots. (B) SCS lead positioned dorsal to the spinal roots. (C) Spinal cord retracted for visualization to show the SCS leads within the spinal canal.

### Cadaver 1

The first interlaminar contralateral insertion (Attempt C1.3) showed mixed final electrode placement on CT, with 11 of 16 electrodes placed ventrolaterally and 5 electrodes placed dorsally. Dissection confirmed that part of the lead was placed dorsal to the spinal roots (Figure 5B). The second interlaminar contralateral insertion (Attempt C1.4) was successfully placed ventrally, with all 8 electrodes located ventrolateral to the roots according to CT and confirmed by dissection. The sacral hiatus insertion (Attempt C1.5) showed 16 of 16 electrodes placed ventrolaterally on CT, which was confirmed by dissection. The caudal portion of the lead wire, however, wove between two root bundles (Supplementary Figure 2A).

### Cadaver 2

For the first ipsilateral interlaminar insertion (Attempt C2.2), all 8 electrodes were positioned ventrolaterally, confirmed by CT and dissection. The second ipsilateral interlaminar lead (Attempt C2.3) resulted in all 8 electrodes being positioned ventromedially on CT. However, dissection revealed a dural puncture at the site of insertion, and the lead was ultimately placed subdurally (Supplementary Figure 2B). Across all cadavers, this was the only case of unexpected tissue damage. The transforaminal insertion (Attempt C2.6) resulted in ventrolateral placement of all 8 electrodes on CT, which was confirmed by dissection. Insertion through the lower lumbar and sacral levels (C2.1, C2.4, and C2.5) were unsuccessful due to resistance encountered while advancing the leads rostrally. Anatomical dissection revealed spinal stenosis at L5, explaining why the leads could not be advanced.

### Cadaver 3

The first interlaminar contralateral insertion (Attempt C3.1) showed 8 of 8 electrodes placed ventrolaterally on CT, which was confirmed by dissection. The first interlaminar ipsilateral insertion (Attempt C3.4) produced both ventromedial and ventrolateral placement, with 6 of 8 electrodes positioned ventromedially and 2 of 8 positioned ventrolaterally. Dissection confirmed that the lead was ventral to the roots. The second interlaminar ipsilateral insertion (Attempt C3.7) showed a combination of ventromedial and ventrolateral positions as well, with 4 of 8 electrodes placed ventromedially and the remaining 4 electrodes placed ventrolaterally. Again, dissection confirmed that the lead was positioned ventral to the roots.

## Discussion

### Clinical Feasibility

We have demonstrated that SCS leads can be placed ventrally using several modified percutaneous implant techniques under fluoroscopic guidance. The four approaches tested here all proved to be viable implant techniques, with nearly all electrodes placed ventrally. Each method, however, presents distinct advantages and limitations, and ultimately, it is at the provider’s discretion to decide which technique would be optimal for each patient. Our data suggest that the sacral hiatus, transforaminal, and interlaminar contralateral approaches lead to reliable ventrolateral placement, whereas the interlaminar ipsilateral approach had mixed ventromedial and ventrolateral placement.

The sacral hiatus approach provided immediate ventral access without navigating around the thecal sac. However, such a caudal starting point may limit maneuverability when targeting more rostral locations. The transforaminal approach also provided direct access to the ventral epidural space, though we advise that practitioners assess for foraminal narrowing with pre-procedural 3D imaging to avoid complications or injury. This approach could be further facilitated by an endoscope to provide visualization of the roots upon entry through the intervertebral foramen. The interlaminar approaches were the most similar to conventional SCS_D_ implant techniques and may feel more familiar to clinicians. Both contralateral and ipsilateral interlaminar approaches proved to be viable, but the ipsilateral technique led to more reliable steering toward midline, while the contralateral approach tended to result in slightly more lateral placements, with some electrodes placed dorsally. Notably, our physician team reported greater difficulty navigating towards midline with the interlaminar contralateral technique. Based on these findings, we suggest that all four approaches are technically feasible, and that optimal technique selection may depend on patient anatomy, physician preference, and desired location in the ventral epidural space.

The target location within the ventral epidural space may depend on the intended therapeutic goal. Ventrolateral placement may be preferable when unilateral muscle activation is desired, allowing the legs to be targeted independently for movements such as cycling or walking. Ventromedial placement may be more likely to activate muscles bilaterally. This bilateral activation could be advantageous for activities that require the same muscles on both legs to be active at the same time, such as standing. However, ventromedial placement may reduce the overall specificity of muscle activation if the leg muscles cannot be activated independently.

### Pre-Operative Considerations

As with any percutaneous spinal intervention, the technical feasibility of ventral access must be considered in the context of patient-specific anatomy. Patient-specific spinal anatomy, especially degenerative changes such as stenosis or arthritis, will influence which technique may be most effective (22). The stenosis encountered in Cadaver 2 illustrates how such anatomical abnormalities can impede lead maneuverability, demonstrating the importance of preoperative imaging for implantation approach selection, such as magnetic resonance imaging (MRI).

Preoperative MRI is routinely obtained prior to SCS implantation to evaluate spinal canal patency and guide procedural planning (23), and may help avoid technical challenges such as those encountered in Cadaver 2. These considerations may be especially relevant in people with chronic SCI, who can exhibit accelerated degenerative changes of the spine over time (24). In addition, many individuals with chronic SCI have prior spinal instrumentation or develop osseous regrowth following decompressive procedures such as laminectomy, which may complicate electrode steering and placement (25).

### Comparison of Dorsal and Ventral SCS

Traditional SCS_D_ primarily activates large-diameter afferent fibers within the dorsal root and dorsal horn (8). These afferents indirectly recruit motoneurons through spinal reflex pathways, producing broad and relatively non-specific muscle activation patterns (9,18). In contrast, SCS_V_ may activate motoneurons in the ventral horn and roots more directly, enabling more selective and controllable muscle activation (21). Consistent with this hypothesis, computational modeling studies of the rat spinal cord have shown that SCS_V_ produces more selective muscle activation than SCS_D_ (20).

SCS_D_ has also been shown to enhance volitional motor control in individuals with chronic paralysis (11,15–17). A proposed mechanism for this functional recovery is that SCS_D_ induces synchronized afferent activity, and residual supraspinal input may gate motoneuron recruitment in response to this synchronized input (26). Because SCS_V_ may primarily activate motoneurons rather than afferent-mediated spinal circuitry, it may not confer the same rehabilitative effects on voluntary control. Instead, SCS_V_ may be better suited for neuroprosthetic applications requiring direct and controllable motor output, such as brain–spine (digital bridge) systems (27). However, it is possible that SCS_V_ could activate afferent fibers near the motoneurons, similar to intraspinal microstimulation, since axons are more readily excitable by electric fields than somas (28). In this case, SCS_V_ could lead to rehabilitative effects with greater specificity.

### Future Work

Before SCS_V_ can be broadly adopted as a therapy or deployed as a neuroprosthetic interface, further studies are needed to establish tolerability of both the ventral lead implant procedure and the delivery of stimulation. Although the current study demonstrates the feasibility of ventral epidural access, it remains unclear whether the implant approaches tested here can be performed in live patients without causing discomfort. However, several case studies and case series have shown that SCS_V_ can treat intractable chronic pain (29–32), with more recent works demonstrating the feasibility of accessing the ventral epidural space using percutaneous leads in live patients (29,30). While these findings are promising, larger and more comprehensive studies are needed to further determine the feasibility and tolerability of SCS_V_ lead placement. Additionally, future work must evaluate whether SCS_V_ can consistently achieve greater muscle specificity than SCS_D_ in humans, as suggested by computational modeling (20). Another important consideration is that direct motoneuron recruitment may produce more rapid muscle fatigue compared to afferent-mediated motoneuron activation (33). Therefore, muscle fatiguability with SCS_V_ must be determined and compared to SCS_D_ and functional electrical stimulation. Collectively, these assessments are necessary to define the therapeutic scope, safety, and long-term viability of SCS_V_ prior to clinical translation.

With finer control of movement via SCS_V_, it will likely be necessary to develop control strategies to deliver stimulation that activates the appropriate muscles at the correct time during walking or other locomotor tasks. Some customization of SCS_D_ control has been demonstrated, particularly in changing between modes for standing, walking, and other activities such as cycling or swimming (16). Notably, there has been significant progress in developing adaptive and personalized control strategies for other stimulation methods, including intraspinal microstimulation and functional electrical stimulation (34–37). These methods could be tailored to SCS_V_ to provide refined and adaptable motor control in people with paralysis.

### Limitations

A limitation of this study is the absence of cerebrospinal fluid (CSF) in cadaver models, which may result in implantation mechanics that differ from those in live patients. In the cadaveric specimens, the lack of CSF eliminated normal distension of the dura mater, and may have enlarged the epidural space (38). Together, these factors, along with the greater friability of the cadaveric tissue, likely increased the susceptibility of the dura mater to puncture during interlaminar SCS implantation Attempt C2.3. Dural puncture is a known but rare procedural risk for percutaneous SCS implants (39). In live tissue, the incidence of dural puncture during interlaminar SCS implantation is approximately 0.48% (40). Standard precautions to reduce this risk include use of the loss-of-resistance technique (41), which was not performed for every implant in this cadaveric study but would be routinely performed in human procedures. Additionally, in the event of a dural puncture in live patients, CSF exiting through the Tuohy needle during lead or stylet advancement would provide immediate feedback to the provider. As a result, dural puncture is both less likely to occur and more readily detectable in live humans, reducing the risk of tissue injury during lead advancement.

## Conclusions

Four different approaches to implanting percutaneous SCS leads along the ventral aspect of the thoracolumbar spinal cord were proposed and compared. All four approaches were successful in placing the leads ventrally, with varying degrees of complexity and likelihood for ease of translation. Taken together, these findings suggest that ventral placement of SCS leads may provide a new interface for enhancing movement after paralysis. If SCS_V_ enables more selective activation of lower-limb muscles, it could support closed-loop neuroprosthetic systems and enhance motor rehabilitation outcomes beyond what is achievable with SCS_D_ alone. While substantial work remains, establishing the feasibility of SCS_V_ implantation represents an important step toward expanding therapeutic options for individuals living with SCI and other motor disorders.

## Data Availability

All data produced in the present study are available upon reasonable request to the authors

## Acknowledgments

The authors would like to thank the Department of Physical Medicine and Rehabilitation at the University of Utah for providing pilot grant funding to support this work. Additionally, we thank Nevro for donating the majority of the spinal cord stimulator leads used in this study. We would also like to acknowledge the staff of the Surgical Knowledge Integration (SKI) Lab at the University of Utah for their help in facilitating the implantation procedures.

---

**Supplementary Figure 1.**
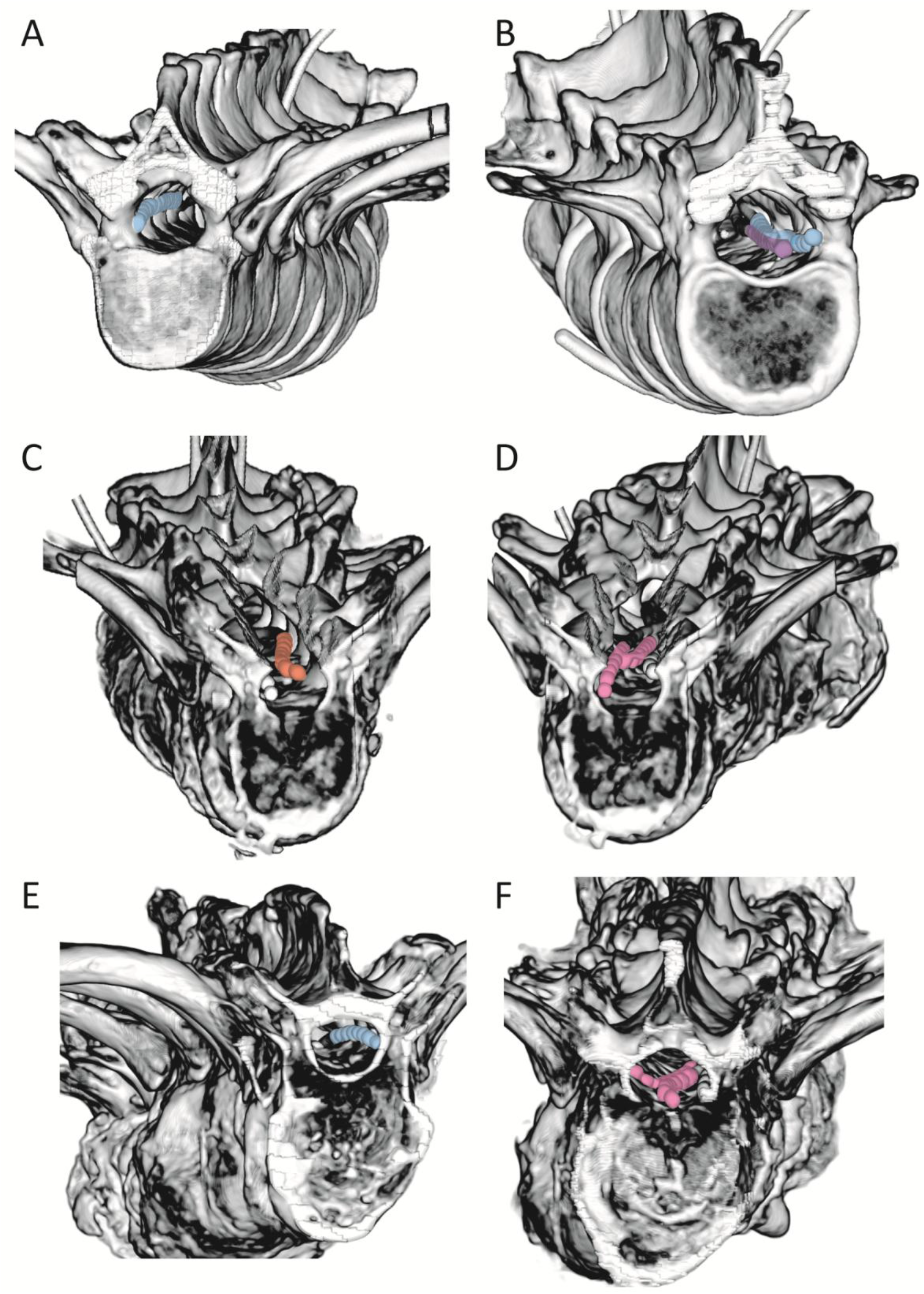
Visualization of SCS leads within the spinal canal from three-dimensional reconstructions of CT scans. Panels (A) and (B) show lead placement for Cadaver 1, panels (C) and (D) show lead placement for Cadaver 2, and panels (E) and (F) show lead placement for Cadaver 3. Lead color indicates implantation method: sacral hiatus (purple), transforaminal (orange), interlaminar contralateral (blue), and interlaminar ipsilateral (pink).

**Supplementary Figure 2.**
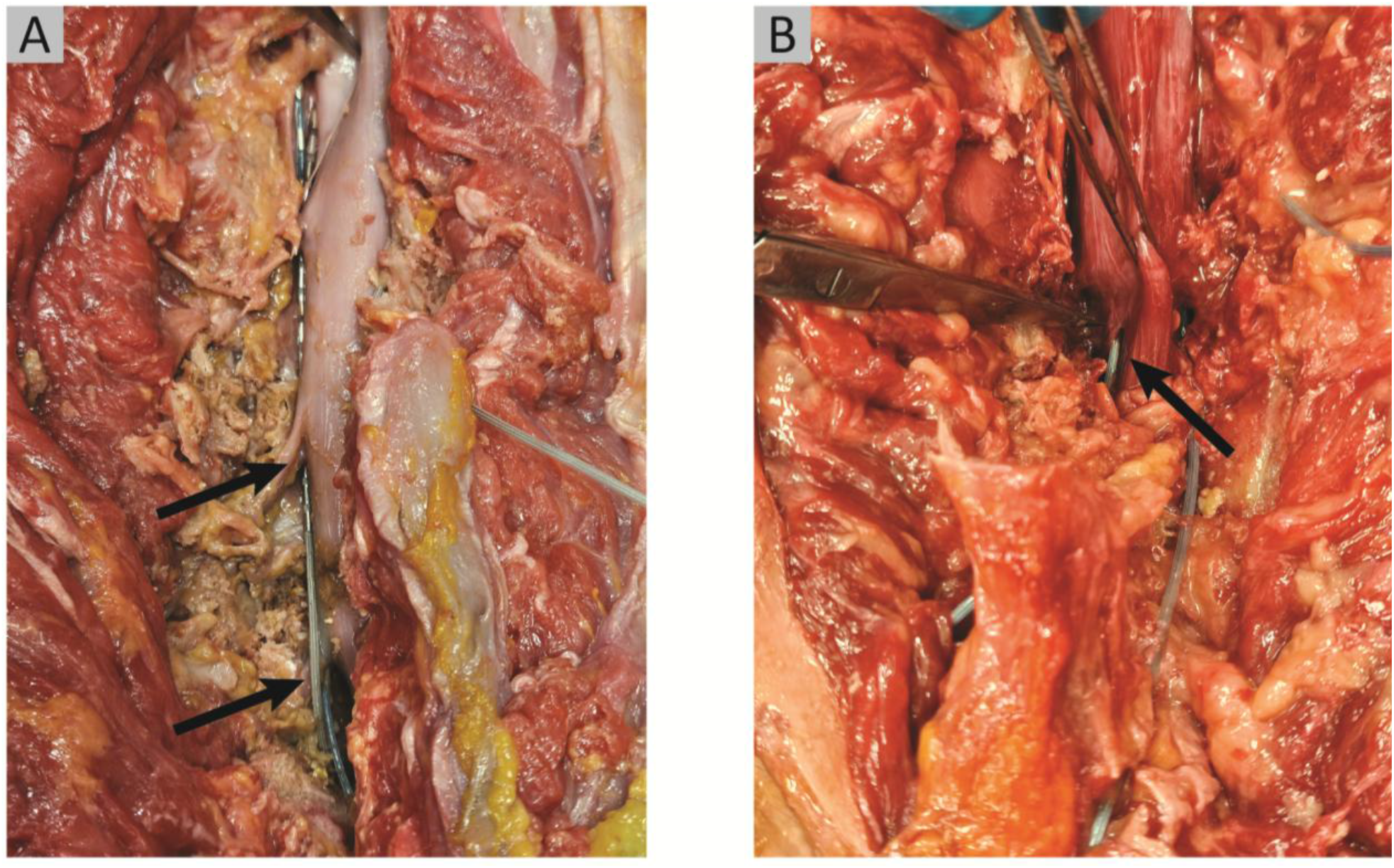
Dissection images of unexpected findings, indicated with the arrows. (A) During implant C1.5, the SCS lead wove between the roots as it was advanced rostrally, with part of the lead wire placed dorsally over the caudal root and ventrally under the rostral root. (B) Dura puncture occurred during implant attempt C2.3.

